# Circadian Disruption Induces Testosterone Decline and Male Reproductive Impairment: Evidence from Epidemiological Studies and Animal Experiments

**DOI:** 10.64898/2026.05.14.26353270

**Authors:** Junxiang Chen, Yangyang Xu, Min Zhao, Jing Liao, Yifei Liu, Yanxi Zhuo, Huihong Cai, Yingping Cao, Heqing Shen, Yu Jiang, Jing Li

## Abstract

This study aims to elucidate the association of circadian rhythm disruption with male testosterone levels and reproductive health using integrated epidemiological and experimental evidence. In the UK Biobank (n = 38,562), rest-activity rhythm amplitude was associated with lower serum testosterone levels (−0.21 nmol/L comparing the lowest vs. highest quartiles) and increased risks of orchitis and hydrocele (hazard ratios: 1.23 and 1.14, respectively). These findings were replicated in an occupational study of shift workers in China (n = 118), where shift work was independently associated with decreased testosterone levels (β = −0.301, *P* = 0.015). In mouse models, circadian disruption induced testicular and epididymal atrophy, spermatogenic disorders, and suppressed circulating testosterone levels, accompanied by downregulation of key steroidogenic proteins. Together, these findings provide converging evidence that circadian rhythm disruption impairs testosterone synthesis, potentially through dysregulation of steroidogenesis, highlighting circadian rhythm as a modifiable environmental determinant of male reproductive health.

**Synopsis:** Modern lifestyle-related environmental stressors (e.g., artificial light at night, jet lag, shift work) disrupt intrinsic circadian rhythms. This circadian misalignment significantly impairs male reproductive health by lowering testosterone levels and compromising testicular and epididymal function.

## Introduction

Male reproductive health is currently facing severe challenges worldwide, with growing concern over reproductive endocrine dysfunction and male reproductive disorders. Among these, low testosterone levels have been identified as a central contributor to male reproductive impairment ^1,2^. Accumulating epidemiological evidence suggests that testosterone levels have shown a widespread decline across populations, with an alarming increase in testosterone deficiency even among younger men ^3,4,5^. Testosterone secretion is regulated by multiple factors, including genetics, age, environmental exposure and unhealthy lifestyles ^6,7,8,9^. In modern societies, however, environmental and behavioral determinants appear to play an increasingly prominent role. Against this backdrop, circadian rhythm disruption (CRD) has emerged as a pervasive and potentially important risk factor for endocrine and reproductive health.

CRD, characterized by misalignment between endogenous biological rhythms and the external light-dark cycle, has become common in contemporary society due to artificial light exposure at night, chronic sleep deprivation, and shift work ^10,11^. It is estimated that up to 50% of the population in industrialized societies experience misalignment between their biological clock and daily routines ^12^. Previous studies have linked CRD to a wide range of adverse health outcomes, including metabolic, cardiovascular, and neurobehavioral disorders ^13,14,15^. However, its specific role in male reproductive health, particularly in testosterone regulation, remains insufficiently characterized.

As the principal male sex hormone, testosterone is essential for spermatogenesis, reproductive organ development, and the maintenance of male secondary sexual characteristics. Notably, testosterone secretion is itself under circadian control, exhibiting a clear diurnal rhythm that typically peaks in the early morning and declines to its lowest levels at night ^16,17^. This temporal pattern suggests that disruption of circadian organization may directly interfere with testosterone homeostasis. In addition to rhythmic secretion, testosterone production depends on tightly coordinated steroidogenic processes within the testes. Key molecules involved in this pathway include steroidogenic acute regulatory protein (StAR), which mediates cholesterol transport into mitochondria as the rate-limiting step of steroidogenesis, as well as 3β-hydroxysteroid dehydrogenase 1 (HSD3B1) and 17α-hydroxylase/17,20-lyase (CYP17A1), which catalyze critical enzymatic conversions in testosterone biosynthesis ^18,19^. CYP19A1 further contributes to androgen–estrogen balance through aromatization ^18,19^. Dysregulation of these steroidogenic pathways may impair testosterone synthesis and contribute to hypogonadism, defective spermatogenesis, reduced sperm quality, and infertility ^20,21,22^. To date, studies have shown StAR protein expression in testicular Leydig cells ^23,24^; environmental endocrine disruptors such as bisphenol A inhibit the mRNA and protein expression of key enzymes, including CYP11A1 and CYP17A1 ^25^; sedentary behavior and insufficient sleep may also impair testicular endocrine function through indirect pathways ^26^. Despite the established role of these enzymatic pathways in testosterone regulation, it remains unclear whether CRD exerts a direct adverse effect on testosterone synthesis beyond disturbing its circadian secretion pattern.

To address these gaps, this study employed a multi-level research approach integrating population-based, occupational, and experimental evidence. First, using data from the UK Biobank, we examined the associations between accelerometer-derived circadian rhythmicity and testosterone levels, as well as male reproductive outcomes. Second, we conducted an occupational study among shift workers in Fuzhou, China, to validate the observed associations in a real-world working population. Third, we employed a mouse model of chronic circadian rhythm disruption to explore pathological and molecular changes in the male reproductive system. By combining epidemiological and experimental evidence, this study aimed to provide a more comprehensive understanding of the role of CRD in testosterone decline and male reproductive impairment, and to offer stronger evidence for the prevention of circadian-related reproductive health damage.

## Methods

### UK Biobank Study

#### Study population

The UK Biobank is a large prospective study that recruited over 500,000 middle-aged participants from 22 assessment centers across England, Scotland, and Wales between 2006 and 2010 ^27^. At recruitment, participants completed questionnaires and interviews, underwent standard physical measurements, and provided biological samples for biomarker measurement. Between 2013 and 2015, a subset of over 100,000 participants underwent 7-day physical activity monitoring using a wrist-worn accelerometer. In the present study, 38,562 male participants with valid accelerometer data were included after excluding those without acceleration intensity time-series data, with low accelerometer data quality, and without measurement of circulating testosterone (**Supplementary Figure 1**).

#### Assessment of circadian rhythmicity

Circadian rhythmicity was assessed using data from the AX3 triaxial accelerometer (Axivity, Newcastle upon Tyne, UK), which participants wore continuously on the dominant wrist for 7 consecutive days during usual daily activities. Raw accelerometer data were processed by the UK Biobank accelerometer expert working group to generate physical activity intensity data in 5-s epochs ^28^.

The relative amplitude of the rest-activity rhythm was used as the indicator of circadian rhythmicity ^29,30^. Relative amplitude was defined as the difference between the most active continuous 10-h period (M10) and the least active continuous 5-h period (L5) within an average 24-h cycle (midnight to midnight), calculated as:

Relative amplitude = (M10-L5)/(M10+L5)

To derive this measure, minute-level averages were first calculated from the 5-s accelerometer data across the 7-day monitoring period. Moving averages were then computed across all possible 5-h and 10-h windows within a 24-h period. The maximum value among the 10-h averages was defined as M10, and the minimum value among the 5-h averages was defined as L5. Relative amplitude ranges from 0 to 1, with lower values indicating greater disruption of circadian rhythmicity, potentially reflecting reduced daytime activity, increased nighttime activity, or both.

#### Assessment of outcomes

The prespecified outcomes in the UK Biobank analysis were serum testosterone concentrations and incident male reproductive disorders, including infertility, orchitis/epididymitis, and hydrocele/spermatocele. Infertility was excluded from the final analyses because only one incident case was identified in this predominantly older cohort.

Serum testosterone concentrations were measured in baseline serum samples using a competitive binding chemiluminescent immunoassay on the Beckman Coulter DXI 800 platform. Incident reproductive disorders were identified using International Classification of Diseases, Tenth Revision (ICD-10) codes through linkage to primary care records, hospital admissions, and death registry data, including infertility (N46), orchitis/epididymitis (N45), and hydrocele/spermatocele (N43). Follow-up were available up to 1st July 2024 for Scotland, 1st August 2024 for Wales, and 1st September 2024 for England in the present study. Participants with prevalent disease at baseline were excluded from the corresponding incident disease analyses.

### Occupational Study of Shift Workers in Fuzhou

#### Study Participants

This occupational study was approved by the Ethics Committee of Fujian Medical University (Approval No.2022-18). Based on the distribution of enterprises operating shift-work systems in Fuzhou, enterprises with representative shift-work characteristics were selected using stratified random sampling according to enterprise size and industry type. Male employees from the selected enterprises were recruited, and onsite investigation and sample collection were conducted from June to August 2024.

The inclusion criteria for the shift-work population were as follows: male sex, age 18-35 years ^31^, employment tenure of at least 1 year, and provision of written informed consent. The exclusion criteria were reproductive disorders such as orchitis or epididymitis, major endocrine diseases such as hypogonadism or thyroid dysfunction, prior surgery involving the testis, adrenal gland, or pituitary gland; and use within the previous 3 months of androgen therapy, anti-androgen therapy, or medications that may substantially affect testosterone levels, such as glucocorticoids, opioid analgesics, or certain psychotropic drugs ^32^.

After screening according to the above criteria, 158 participants were enrolled. After excluding individuals with incomplete questionnaires or inadequate blood samples, 118 participants were included in the final analysis, comprising 86 shift workers and 32 non-shift workers (**Supplementary Figure 2**).

#### Questionnaire assessment

All investigators received standardized training before data collection. Information was obtained using a structured questionnaire covering demographic characteristics, including age, education level, marital status, and body mass index (BMI), as well as lifestyle factors such as smoking and alcohol consumption.

Shift work was defined as any work schedule deviating from standard daytime working hours (7:00/8:00 to 17:00/18:00), including fixed night shifts and rotating shifts ^33^. Participants who had engaged in such schedules at least 5 times per month during the 6 months preceding the survey were classified as shift workers. Shift work duration was defined as the cumulative number of years since the participant began shift work. Employment tenure was defined as the cumulative number of years under contract with the current employer.

#### Measurement of serum testosterone

Fasting blood samples were collected from all 118 participants in the early morning (8:00-10:00) ^34^. Blood was drawn into anticoagulant-free centrifuge tubes and centrifuged at 3000×g for 10 minutes at 4℃ to separate the serum, which was then stored at -80℃ until detection. Serum testosterone levels were measured by chemiluminescence immunoassay using a Beckman DxI800 automated chemiluminescence immunoanalyzer. All detection procedures were conducted in strict accordance with the instrument operation specifications and laboratory standard protocols to ensure measurement accuracy and repeatability.

### Animal Experiments

#### Animals and circadian disruption model

Thirty-six healthy adult male C57BL/6 mice aged 6-8 weeks were purchased from Shanghai SLAC Laboratory Animal Co., Ltd. Mice were housed in a specific-pathogen-free environment at 22±2°C and 50±5% relative humidity, with free access to standard chow and sterile water. Before the intervention, all animals were acclimatized for 2 weeks under a standard 12-hour light/12-hour dark (LD) cycle (lights on at 07:00 and lights off at 19:00). After acclimatization, mice were housed in rhythm-controlled chambers and randomly assigned to three groups (n=12 per group) using a random number table: a control LD group maintained under a standard 12-hour light/12-hour dark cycle throughout the experiment; a phase-advance (AD) group exposed to a 6-hour weekly advance of the light-dark cycle; and a light-dark reversal (LD-DL) group exposed to a 12-hour weekly reversal of the light-dark cycle. The intervention lasted for 15 consecutive weeks to induce chronic circadian disruption. (**Supplementary Figure 3**). All animal experiments were approved by the Institutional Animal Care and Use Committee (IACUC) and the Animal Experimental Ethics Committee of Fujian Medical University (Approval No.: IACUC FJMU 2022-NSFC-0325).

#### Western blot analysis

Testicular tissues were lysed on ice in RIPA buffer (Solarbio, Beijing, China) supplemented with protease and phosphatase inhibitor cocktails (MedChemExpress, Shanghai, China). Lysates were centrifuged at 13,000×g for 20 minutes at 4°C, and the supernatants were collected for total protein extraction. Equal amounts of protein (40 μg per sample) were denatured in 5×SDS-PAGE loading buffer (Epizyme, China) by boiling for 10 minutes. Proteins were separated by 8%-12% SDS-PAGE and transferred onto PVDF membranes (Millipore, USA). Membranes were blocked with 5% non-fat milk in TBST for 1 hour at room temperature and then incubated overnight at 4°C with primary antibodies against GAPDH, HSD3B1, STAR, CYP17A1, and CYP19A1. After washing, membranes were incubated with HRP-conjugated secondary antibodies (Thermo Fisher Scientific, USA) at a dilution of 1:5000 for 1 hour at room temperature. Protein bands were visualized using an ECL detection system (Tanon, China), and band intensities were quantified using ImageJ software. Target protein expression was normalized to GAPDH.

#### H&E staining

Testes and epididymides were fixed in tissue fixative (Servicebio, Wuhan, China) for 24 hours at room temperature. After fixation, tissues were dehydrated through graded ethanol, cleared in xylene, embedded in paraffin, and sectioned at a thickness of 5 μm. Sections were rehydrated through graded ethanol and stained with hematoxylin and eosin according to standard procedures. After staining, sections were dehydrated, cleared, mounted with neutral balsam, and examined under a light microscope for histopathological evaluation.

#### Measurement of mouse serum testosterone

Mouse serum testosterone concentrations were measured using the Elabscience QuicKey Pro Mouse Testosterone ELISA Kit (Cat. No. E-OSEL-M0003). Serum samples were centrifuged at 3,000×g for 10 minutes at 4°C before analysis. Standards, samples, and blank controls were added to duplicate wells of the pre-coated 96-well plate. Absorbance was measured at 450 nm with 630 nm as the reference wavelength within 10 minutes using a microplate reader. Testosterone concentrations were calculated from a 4-parameter logistic standard curve (R²>0.99) generated in GraphPad Prism 9.

### Statistical methods

For the UK Biobank analysis, participant characteristics were summarized as means with standard deviations (SD) for continuous variables and counts with percentages (%) for categorical variables. Differences across quartiles of relative amplitude were tested using one-way ANOVA analysis for continuous variables and chi-squared test for categorical variables. Generalized linear regression models were used to estimate the association between relative amplitude and circulating testosterone concentrations, and Cox proportional hazard regression models were used to estimate the hazard ratios (HRs) for incident reproductive disorders. Model 1 was adjusted for age at accelerometry. Model 2 was additionally adjusted for ethnicity, education, Townsend deprivation index, BMI category, smoking status, alcohol consumption, healthy diet, and season at accelerometry.

For the occupational study, differences between shift workers and non-shift workers were evaluated using the independent-sample*s t-test* for continuous variables and the chi-squared test for categorical variables. Multiple linear regression was used to examine the independent association between shift work and serum testosterone concentration, with testosterone concentration as the dependent variable and shift-work status as the primary independent variable. Age, education level, marital status, smoking status, alcohol consumption, BMI category, and employment tenure were included as covariates. Multicollinearity was assessed using the variance inflation factor (VIF), with VIF > 10 indicating substantial collinearity.

For the animal experiments, comparisons among the three groups were performed using one-way ANOVA analysis of variance followed by the LSD post hoc test.

Statistical analyses were performed using SPSS 26.0 and R 4.5.1. A two-sided *P* < 0.05 was considered statistically significant.

## Results

### Participant characteristics in the UK Biobank

At baseline, the participants had a mean (SD) age of 56.80 (7.89) years and a TDI of -1.78 (2.81). The majority of the participants were White (96.7%), overweight (49.2%), never smokers (51.9%), frequent drinkers (56.7%), had an educational level of less than college (54.9%), and unhealthy diet (85.6%). Across quartiles of relative amplitude, participants with lower relative amplitude tended to be older, more likely to be of non-White ethnicity, obese, and current smokers, and had a higher Townsend deprivation index. They were also less likely to be frequent drinkers, had lower educational attainment, and were less likely to follow a healthy diet. (**Table 1**).

**Table 1.**
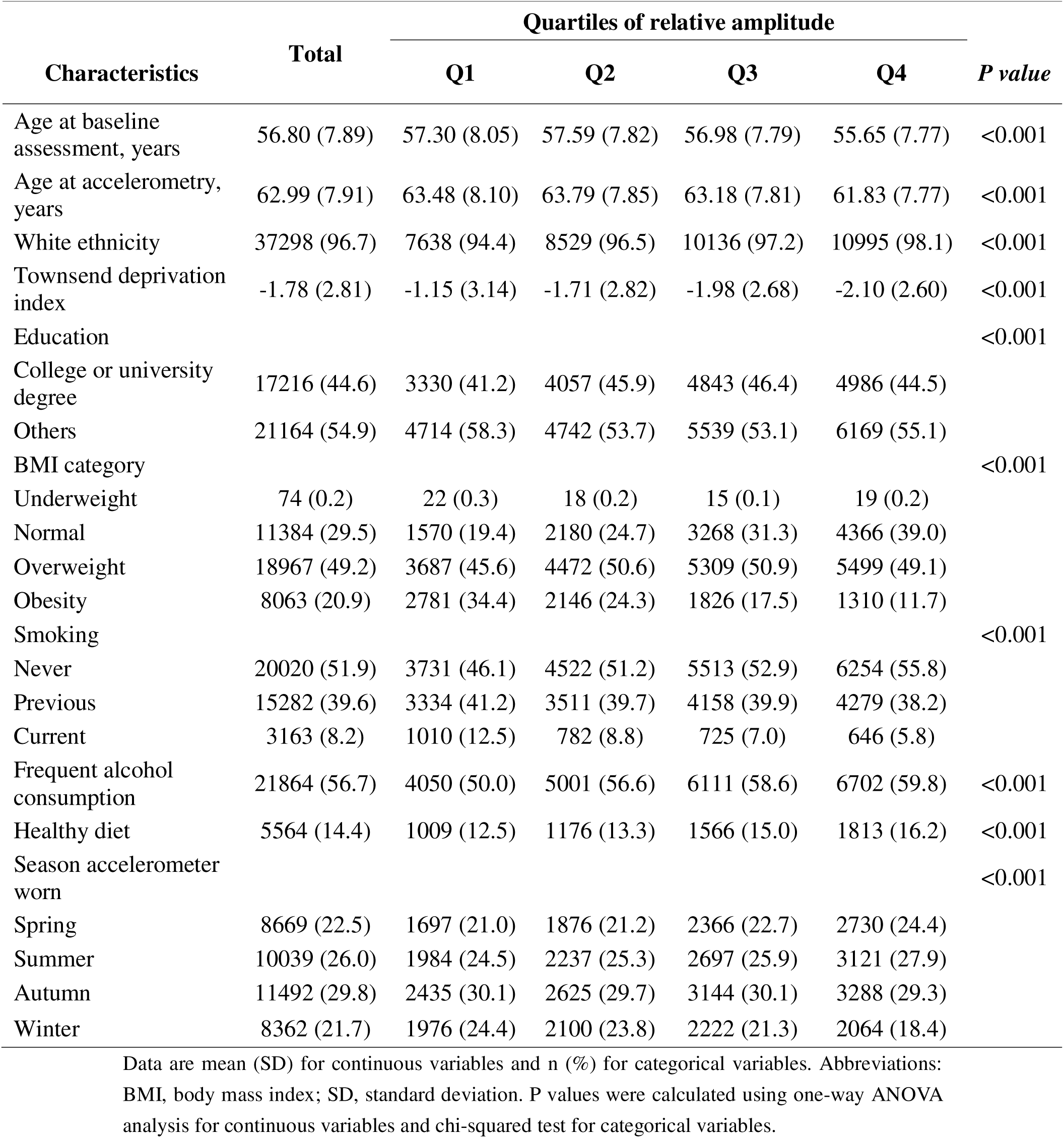
Demographic characteristics of participants by circadian relative amplitude.

### Relative amplitude and testosterone concentration in the UK Biobank

Testosterone concentrations were lower among participants with lower relative amplitude which indicates more disrupted circadian rhythmicity. Mean testosterone levels were 12.43, 12.16, 11.89, and 11.69 nmol/L from the highest to the lowest quartile of relative amplitude, respectively (**Table 2**, **Figure 1**). In the age-adjusted model, men in the lowest quartile had substantially lower testosterone concentrations than those in the highest quartile. After multivariable adjustment in Model 2, the association remained statistically significant. Compared with participants in the highest quartile, those in the lowest quartile had 0.21 nmol/L (95%CI: 0.11, 0.32) lower testosterone concentrations. In addition, each 1-SD decrease in relative amplitude was associated with a 0.07 nmol/L (95%CI: 0.03, 0.10) reduction in testosterone concentration (95%CI: −0.10, −0.03).

**Figure 1.**
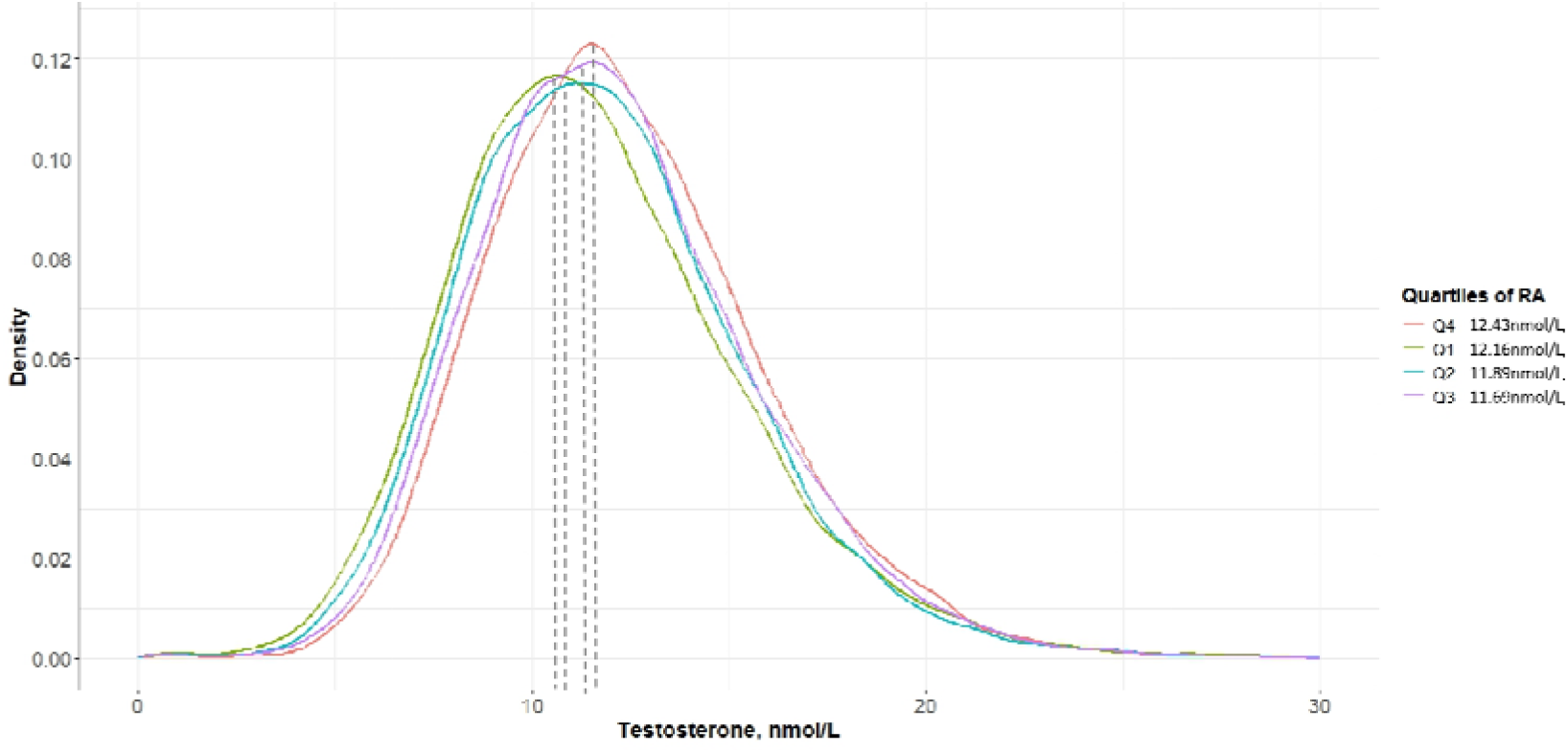
Testosterone concentrations by quartiles of relative amplitude.

**Table 2.**
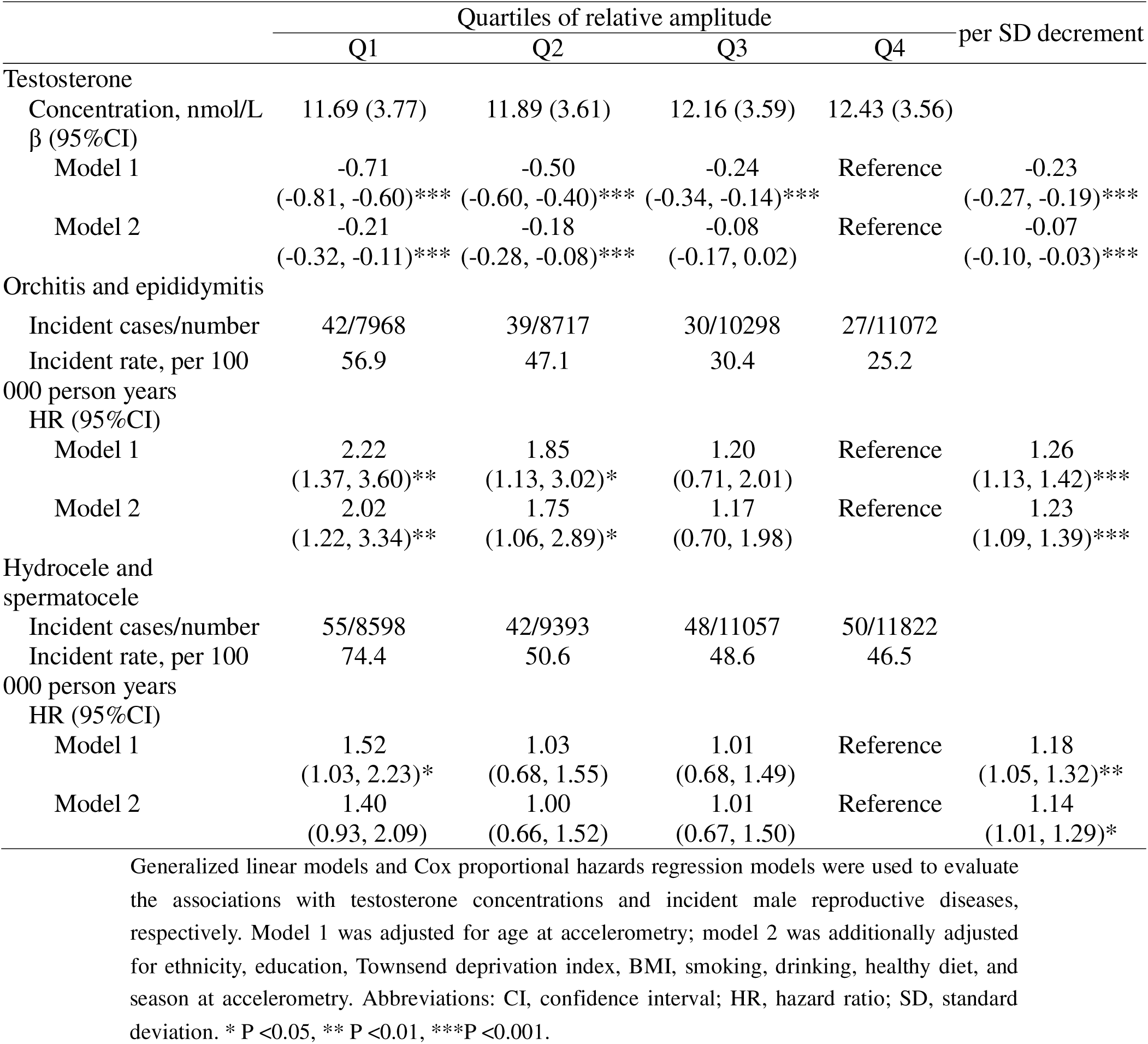
Associations of relative amplitude with the concentration of testosterone, and the risk of male reproductive diseases.

|  | Quartiles of relative amplitude |  |  |  | per SD decrement |
| --- | --- | --- | --- | --- | --- |
|  | Q1 | Q2 | Q3 | Q4 |  |
| **Testosterone** |  |  |  |  |  |
| Concentration, nmol/L | 11.69 (3.77) | 11.89 (3.61) | 12.16 (3.59) | 12.43 (3.56) |  |
| $\beta$ (95%CI) | | | | | |
| Model 1 | -0.71<br>(-0.81, -0.60)*** | -0.50<br>(-0.60, -0.40)*** | -0.24<br>(-0.34, -0.14)*** | Reference | -0.23<br>(-0.27, -0.19)*** |
| Model 2 | -0.21<br>(-0.32, -0.11)*** | -0.18<br>(-0.28, -0.08)*** | -0.08<br>(-0.17, 0.02) | Reference | -0.07<br>(-0.10, -0.03)*** |
| **Orchitis and epididymitis** |  |  |  |  |  |
| Incident cases/number | 42/7968 | 39/8717 | 30/10298 | 27/11072 |  |
| Incident rate, per 100<br>000 person years | 56.9 | 47.1 | 30.4 | 25.2 |  |
| HR (95%CI) |  |  |  |  |  |
| Model 1 | 2.22<br>(1.37, 3.60)** | 1.85<br>(1.13, 3.02)* | 1.20<br>(0.71, 2.01) | Reference | 1.26<br>(1.13, 1.42)*** |
| Model 2 | 2.02<br>(1.22, 3.34)** | 1.75<br>(1.06, 2.89)* | 1.17<br>(0.70, 1.98) | Reference | 1.23<br>(1.09, 1.39)*** |
| **Hydrocele and spermatocele** |  |  |  |  |  |
| Incident cases/number | 55/8598 | 42/9393 | 48/11057 | 50/11822 |  |
| Incident rate, per 100<br>000 person years | 74.4 | 50.6 | 48.6 | 46.5 |  |
| HR (95%CI) |  |  |  |  |  |
| Model 1 | 1.52<br>(1.03, 2.23)* | 1.03<br>(0.68, 1.55) | 1.01<br>(0.68, 1.49) | Reference | 1.18<br>(1.05, 1.32)** |
| Model 2 | 1.40<br>(0.93, 2.09) | 1.00<br>(0.66, 1.52) | 1.01<br>(0.67, 1.50) | Reference | 1.14<br>(1.01, 1.29)* |

Generalized linear models and Cox proportional hazards regression models were used to evaluate the associations with testosterone concentrations and incident male reproductive diseases, respectively. Model 1 was adjusted for age at accelerometry; model 2 was additionally adjusted for ethnicity, education, Townsend deprivation index, BMI, smoking, drinking, healthy diet, and
season at accelerometry. Abbreviations: CI, confidence interval; HR, hazard ratio; SD, standard deviation. \* $P < 0.05$ , \*\* $P < 0.01$ , \*\*\* $P < 0.001$ .

### Relative amplitude and incident male reproductive disorders

Lower relative amplitudes was also associated with a higher incidence of male reproductive disorders. Specifically, in multivariable-adjusted Model 2, each 1-SD decrease in relative amplitude was associated with a 23% higher risk of orchitis/epididymitis (HR=1.23; 95%CI: 1.09, 1.39) and a 14% higher risk of hydrocele/spermatocele (HR=1.14; 95%CI: 1.01, 1.29).

### Characteristics of the occupational study population in Fuzhou

This study included a total of 118 male occupational participants, including 86 shift workers and 32 non-shift workers. Compared with non-shift workers, shift workers were younger and had lower serum testosterone concentrations. The mean (SD) testosterone concentration was 3.8 (1.4) ng/mL in shift workers and 4.6 (1.5) ng/mL in non-shift workers. Significant between-group differences were also observed for age and BMI, whereas educational level, marital status, smoking status, alcohol consumption, and employment tenure did not differ significantly between the two groups (**Table 3**).

**Table 3.** General demographic characteristics of male shift workers in Fuzhou.

| Characteristics | Total<br>(n=118) | Non-Shift<br>(n=32) | Shift<br>(n=86) | *P value* |
| --- | --- | --- | --- | --- |
| **Age (years)** |  | 31.0 (3.3) | 27.4 (3.6) | < 0.05 |
| 21-28 | 66 (55.9) | 7 (21.9) | 59 (68.6) |  |
| 29-35 | 52 (44.1) | 25 (78.1) | 27 (31.4) |  |
| **Educational Level** |  |  |  | 0.08 |
| High school and below | 60 (50.8) | 14 (43.8) | 46 (53.5) |  |
| College or university | 56 (47.5) | 16 (50.0) | 40 (46.5) |  |
| Master's and above | 2 (1.7) | 2 (6.2) | 0 |  |
| **Marital Status** |  |  |  | 0.16 |
| Unmarried | 69 (58.5) | 13 (40.6) | 56 (65.1) |  |
| Married | 49 (41.5) | 19 (59.4) | 30 (34.9) |  |
| **Smoking Status** |  |  |  | 0.30 |
| Never smoked | 62 (52.5) | 21 (65.6) | 41 (47.7) |  |
| <10 cigarettes/day | 19 (16.1) | 2 (6.2) | 17 (19.8) |  |
| 10-20 cigarettes/day | 26 (22.0) | 6 (18.8) | 20 (23.3) |  |
| >20 cigarettes/day | 10 (8.5) | 3 (9.4) | 7 (8.1) |  |
| Quit smoking(> 6 months) | 1 (0.9) | 0 | 1 (3.1) |  |
| **Alcohol Consumption** |  |  |  | 0.85 |
| Never drank | 91 (77.1) | 26 (81.3) | 65 (75.6) |  |
| 1-2 times/week | 21 (17.8) | 5 (15.6) | 16 (18.6) |  |
| 2-3 times/week | 2 (1.7) | 0 | 2 (2.3) |  |
| >3 times/week | 2 (1.7) | 0 | 2 (2.3) |  |
| Quit drinking | 2 (1.7) | 1 (3.1) | 1 (1.2) |  |
| **BMI ( kg/m<sup>2</sup> )** |  |  |  | < 0.05 |
| < 18.5 | 6 (5.1) | 2 (6.2) | 4 (4.6) |  |
| 18.5 ≤ BMI < 24.0 | 52 (44.1) | 12 (37.5) | 40 (46.5) |  |
| 24.0 ≤ BMI < 28.0 | 38 (32.2) | 8 (25.0) | 30 (34.9) |  |
| ≥ 28.0 | 22 (18.6) | 10 (31.3) | 12 (14.0) |  |
| **Employment Tenure (years)** |  | 7.6 (3.7) | 8.6 (2.9) | 0.11 |
| < 5 | 5 (4.2) | 5 (15.6) | 0 |  |
| 5- | 73 (61.9) | 19 (59.4) | 54 (62.8) |  |
| 10- | 33 (28.0) | 7 (21.9) | 26 (30.2) |  |
| ≥ 15 | 7 (5.9) | 1 (3.1) | 6 (7.0) |  |
| **Testosterone level (nmol/L)** |  | 15.8 (5.2) | 13.2 (4.7) | < 0.05 |
| < 10.4 | 25 (21.2) | 4 (12.5) | 21 (24.4) |  |
| ≥10.4 | 93 (78.8) | 28 (87.5) | 65 (75.6) |  |

### Association between Shift Work and Serum Testosterone Levels

In the multivariable linear regression model, shift work was independently associated with lower serum testosterone concentrations. Specifically, shift work was associated with a decrease of 0.968 ng/mL in testosterone concentration (B=−0.968, standardized β=−0.301, *P*=0.015). None of the included covariates showed a statistically significant association with testosterone concentration in this model. Multicollinearity diagnostics indicated acceptable model stability, with all variance inflation factors below 3 (**Table 4**).

**Table 4.** Linear Regression of Shift Work on Serum Testosterone Levels.

|  | Unstandardized Coefficient |  | Standardized Coefficient |  | Significance | Collinearity Statistics |  |
| --- | --- | --- | --- | --- | --- | --- | --- |
|  | B | Standard Error | Beta | t |  | Tolerance | VIF |
| (Constant) | 6.340 | 1.637 |  | 3.873 | <0.001 |  |  |
| Shift work (yes/no) | -0.968 | 0.393 | -0.301 | -2.461 | 0.015 | 0.522 | 1.915 |
| Age | 0.001 | 0.054 | 0.002 | 0.012 | 0.991 | 0.376 | 2.656 |
| Educational level | -0.205 | 0.260 | -0.076 | -0.790 | 0.431 | 0.833 | 1.200 |
| Marital status | -0.459 | 0.316 | -0.158 | -1.454 | 0.149 | 0.661 | 1.514 |
| Smoking status | 0.194 | 0.133 | 0.145 | 1.460 | 0.147 | 0.785 | 1.274 |
| Drinking status | -0.321 | 0.180 | -0.168 | -1.786 | 0.077 | 0.884 | 1.131 |
| BMI | -0.242 | 0.162 | -0.135 | -1.492 | 0.139 | 0.958 | 2.700 |
| Employment Tenure | -0.002 | 0.065 | -0.005 | -0.032 | 0.974 | 0.370 | 2.701 |

### Circadian rhythm disruption induced pathological injury in the testes and epididymis of male mice

To further investigate the biological plausibility of the population findings, we established a chronic circadian disruption model in male C57BL/6 mice. After 15 weeks of light-cycle shifting, testis weight and testis coefficient decreased in both the advance group and the LD-DL group (**Figure 2A, B**). The epididymis weight and epididymis coefficient were also reduced in the advance group (**Figure 2C, D**). Histopathologically, H&E-stained testicular sections showed that the interstitial space between the seminiferous tubules became wider, and the spermatogenic tubules tended to atrophy (**Figure 2E**). The sperm number declined in both the advance group and LD-DL group, as indicated by the H&E staining of the epididymis (**Figure 2F**). These results confirmed that the circadian disruption of male mice led to testicular damage.

**Figure 2.**
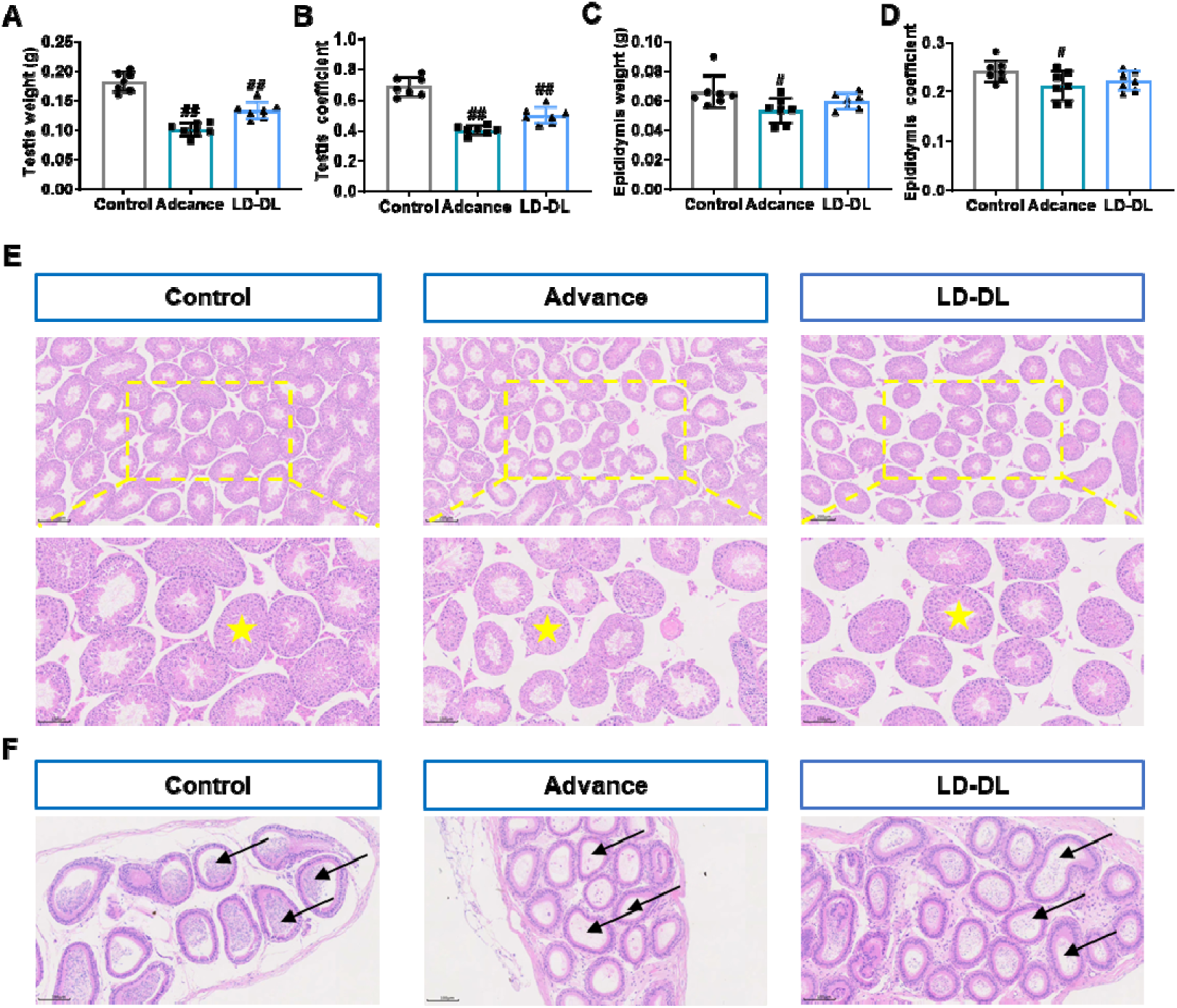
Pathological changes in the testis and epididymis of C57BL/6 mice induced by circadian rhythm disruption. A: Testis weight; B: Testis coefficient; C: Epididymis weight; D: epididymis coefficient; E: H&E staining of mouse testis (Yellow stars: seminiferous tubules), scale bar=200 μm; F: H&E staining of the mouse epididymis (black arrow: epididymal duct), scale bar=100 μm. #: *P*:<0.05, #: *P*<0.01.

### Circadian rhythm disruption reduced testosterone synthesis in male mice

In the mouse model, CRD was also accompanied by a reduction in serum testosterone concentration (**Figure 3A**). Western blot analysis further showed decreased expression of key steroidogenic proteins, including HSD3B1 in both circadian disruption groups, and CYP19A1 and StAR expression in the advance group (**Figure 3B**). The above results demonstrated that CRD induced decreased testosterone *in vivo* model.

**Figure 3.**
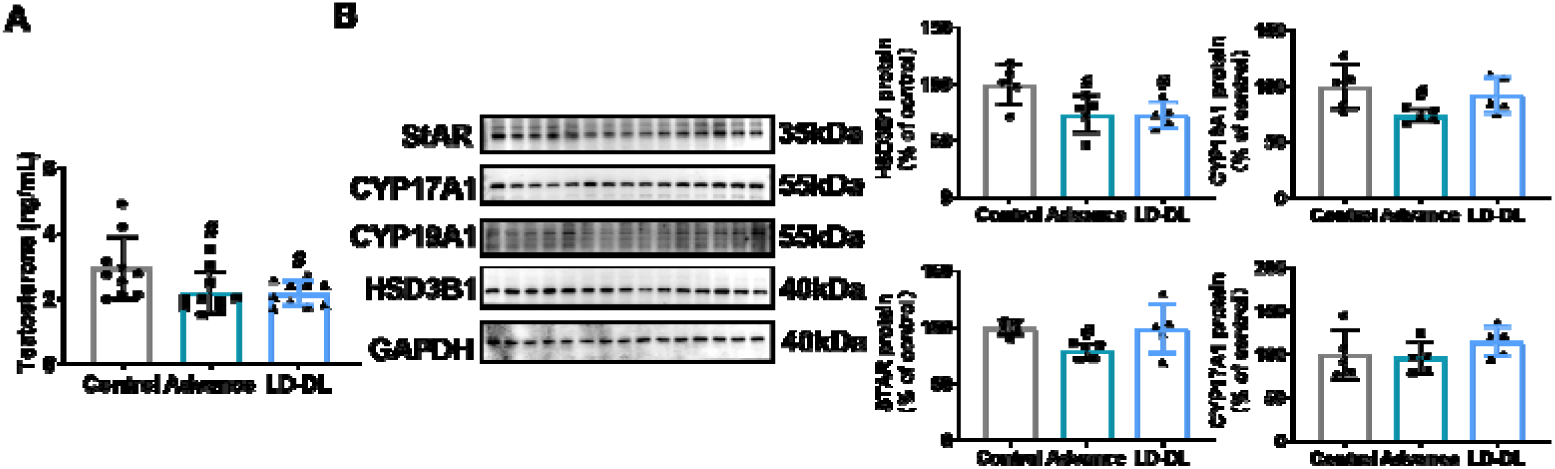
The circadian rhythm of C57BL/6 mice was disrupted, and the testosterone level was decreased. A: Serum testosterone levels in mice; B: Expression levels of testosterone synthesis-related proteins in mouse testes. #: *P*<0.05, ##: *P*<0.01.

## Discussion

In this study, we integrated evidence from a large population-based cohort, an occupational study of young male workers, and a mouse model to examine the relationship between CRD and male reproductive health. Across these complementary approaches, a consistent pattern emerged: disrupted circadian rhythmicity was associated with lower testosterone levels, and experimental circadian disruption induced pathological and molecular changes in the male reproductive system.

In the UK Biobank, lower relative amplitude was associated with lower testosterone concentrations in a dose-dependent manner, even after adjustment for multiple sociodemographic and lifestyle factors. These findings are biologically plausible because testosterone secretion follows a circadian pattern and is sensitive to sleep-wake timing and light-dark alignment ^16,17^. The occupational study provided external support for the population-based findings by showing that shift work was independently associated with lower testosterone concentrations in young men, thereby extending the relevance of the findings to a real-world working population with chronic circadian challenge.

Importantly, the association in the UK Biobank was not limited to testosterone concentrations. Lower relative amplitude was also associated with higher risks of orchitis/epididymitis and hydrocele/spermatocele, suggesting that circadian disruption may have broader implications for male reproductive health beyond endocrine disturbance alone. Taken together with prior studies linking sleep disruption, mistimed light exposure, and shift work to reproductive dysfunction ^35,36,37,38^, our results strengthen the evidence that CRD may represent an independent environmental determinant of male reproductive impairment.

The animal experiments further support this interpretation and provide mechanistic insight. Using a 15-week circadian disruption protocol that mimics recurrent phase shifts and light-dark reversal, we observed reduced circulating testosterone concentrations, lower expression of key steroidogenic proteins, testicular and epididymal atrophy, and histopathological evidence of spermatogenic injury. This model covers a significant proportion of the mouse life cycle ^39^. It can reflect how mild damage turns into long-term harm with only the timing adjusted, and the results are reliable for further application. These findings suggest that CRD may impair testosterone biosynthesis by disrupting steroidogenic machinery, including StAR-dependent cholesterol transport and downstream enzymatic conversion pathways, consistent with the views of recent domestic and foreign studies ^40,41,42^. Although our data do not fully delineate the upstream molecular cascade, they support the hypothesis that circadian disturbance adversely affects Leydig cell endocrine function.

Combined with the reduced testicular coefficient, pathological changes such as widened interstitial space and atrophy of seminiferous tubules, and decreased sperm count in the epididymis observed in animal experiments, our results also suggest that circadian disruption may compromise the spermatogenic microenvironment and increase susceptibility to male reproductive disorders. The spermatogenic microenvironment is critical for spermatogenesis and sperm maturation, and its homeostasis is regulated by the rhythmic expression of core circadian clock genes ^43^. Previous research shows circadian disruption changes core clock genes in testicular Sertoli cells. It harms testis tissue, disturbs sperm cell growth and damages seminiferous tubules, and finally weakens male reproductive function ^44^, which is consistent with our findings. Additionally, the testis is an immune-privileged organ whose immune homeostasis relies on the coordinated function of the blood-testis barrier and Sertoli cells ^45^. We therefore speculate that abnormal clock genes in testicular immune cells may raise the risk of orchitis and epididymitis. These genes boost inflammatory factors and reduce immune tolerance, making the reproductive system easier to get sick ^46,47^. Then, the epididymis, a key site for sperm maturation and storage, is more susceptible to inflammation and immune responses than the testis, for that epididymitis is far more prevalent than isolated orchitis ^48^. Thus, its dysfunction and disrupted spermatogenic microenvironment homeostasis may also raise the risk of male reproductive system diseases. Accordingly, our research supports the possibility that CRD contributes to reproductive injury through combined endocrine, microenvironmental, and potentially inflammatory pathways. These mechanisms warrant further investigation in future studies.

In conclusion, our findings provide converging epidemiological and experimental evidence that circadian disruption is associated with lower testosterone levels and impaired male reproductive health. Maintaining circadian health may therefore represent a potentially important strategy for protecting male reproductive function, particularly in populations chronically exposed to shift work or other forms of circadian misalignment.

## Data Availability

All data produced in the present study are available upon reasonable request to the authors.

## Author Statement

Yu Jiang and Jing Li, as corresponding authors, secured research funding, supervised the research progress, organized experimental work, analyzed data, prepared and revised the manuscript. Heqing Shen conceived and designed the study and revised the manuscript. Junxiang Chen and Yangyang Xu are co-first authors who equally contributed to data collection, experiment implementation and manuscript drafting. Min Zhao, Jing Liao, Yifei Liu, Yanxi Zhuo and Huihong Cai assisted with experiments and data collation. Yingping Cao performed clinical laboratory testing and related data support. All authors have read and approved the final manuscript.

## Acknowledgements and funding

We thank the Public Technology Service Center at Fujian Medical University for giving their technical support in this study. This work was supported by the National Natural Science Foundation of China (grant number: 82204002), Joint Funds for the innovation of science and Technology, Fujian province (Grant number: 2025Y9270, 2024Y9230), Natural Science Foundation of Fujian province (Grant number: 2025J01762), and Start-up funding for high-level talent research of Fujian Medical University (Grant number: XRCZX2024028). The graphical abstract was generated by BioGDP.com ^49^.

---

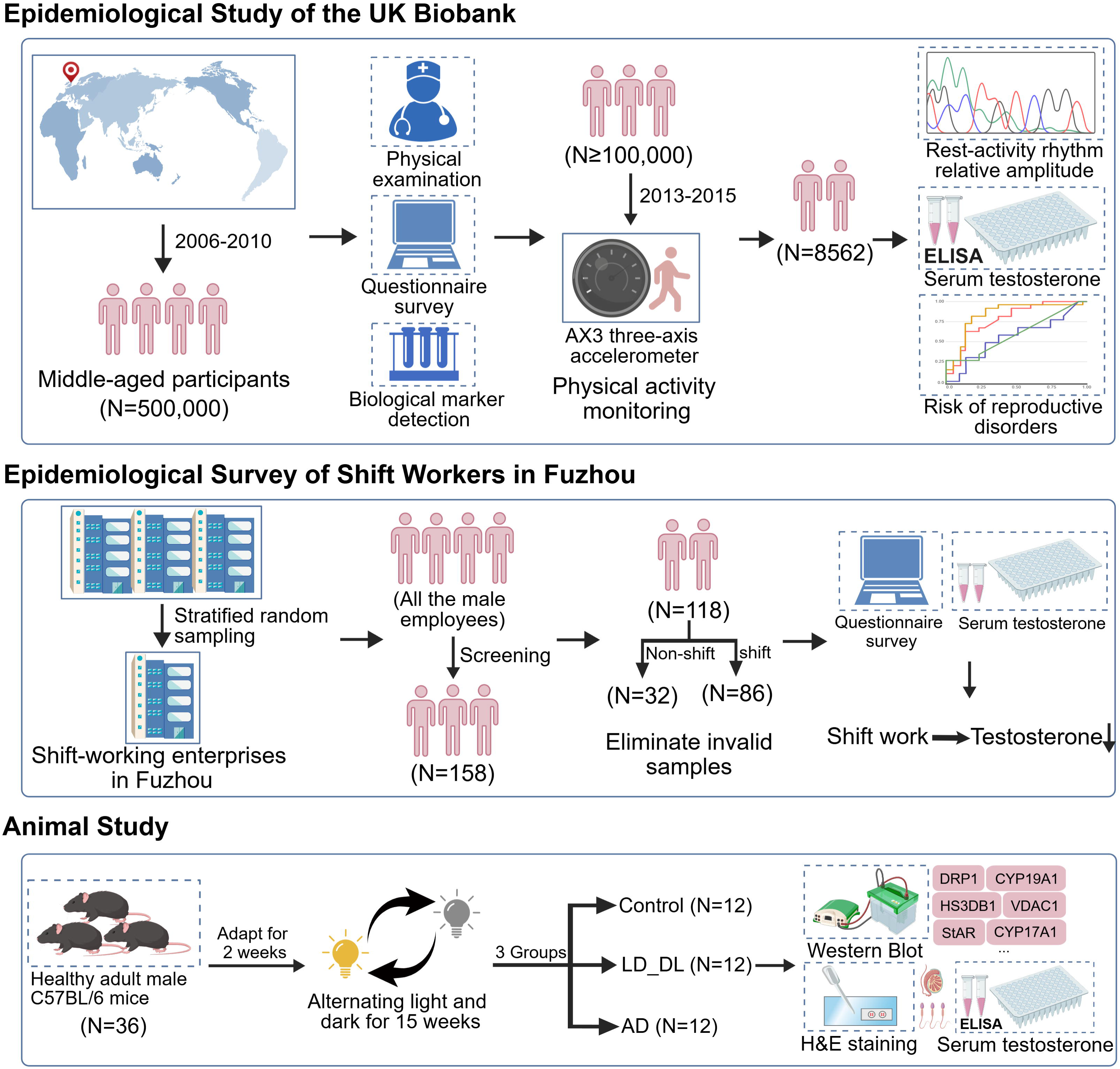

## References

(1) Serbouti, A.; Berrada, K.; Housbane, S.; Louanjli, N.; Aboutaieb, R. Low Testosterone and Sperm Quality Alterations: A Prospective Study of Sperm DNA Fragmentation and Chromatin Condensation in Infertile Men. Biology (Basel) 2026, 15 (3). DOI: 10.3390/biology15030287 From NLM.

(2) Fraile-Martínez, Ó.; Ortega, M. A.; García-Montero, C. Understanding the Secular Decline in Testosterone: Mechanisms, Consequences, and Clinical Perspectives. Int J Mol Sci 2026, 27 (2). DOI: 10.3390/ijms27020692 From NLM.

(3) Chodick, G.; Epstein, S.; Shalev, V. Secular trends in testosterone- findings from a large state-mandate care provider. Reprod Biol Endocrinol 2020, 18 (1), 19. DOI: 10.1186/s12958-020-00575-2 From NLM.

(4) Mazur, A.; Westerman, R.; Mueller, U. Is rising obesity causing a secular (age-independent) decline in testosterone among American men? PLoS One 2013, 8 (10), e76178. DOI: 10.1371/journal.pone.0076178 From NLM.

(5) Cohen, J.; Nassau, D. E.; Patel, P.; Ramasamy, R. Low Testosterone in Adolescents & Young Adults. Front Endocrinol (Lausanne) 2019, 10, 916. DOI: 10.3389/fendo.2019.00916 From NLM.

(6) Zornitzki, T.; Tshori, S.; Shefer, G.; Mingelgrin, S.; Levy, C.; Knobler, H. Seasonal Variation of Testosterone Levels in a Large Cohort of Men. Int J Endocrinol 2022, 2022, 6093092. DOI: 10.1155/2022/6093092 From NLM.

(7) Ohlsson, C.; Wallaschofski, H.; Lunetta, K. L.; Stolk, L.; Perry, J. R.; Koster, A.; Petersen, A. K.; Eriksson, J.; Lehtimäki, T.; Huhtaniemi, I. T.;, et al. Genetic determinants of serum testosterone concentrations in men. PLoS Genet 2011, 7 (10), e1002313. DOI: 10.1371/journal.pgen.1002313 From NLM.

(8) Mazurkiewicz, D.; Gajda, R.; Ambrozik-Haba, J.; Bożek, W.; Ceremuga, M.; Serek, P. Effects of Lifestyle, Diet, and Body Composition on Free Testosterone and Cortisol Levels in Young Men. Nutrients 2025, 17 (23). DOI: 10.3390/nu17233772 From NLM.

(9) Duca, Y.; Aversa, A.; Condorelli, R. A.; Calogero, A. E.; La Vignera, S. Substance Abuse and Male Hypogonadism. J Clin Med 2019, 8 (5). DOI: 10.3390/jcm8050732 From NLM.

(10) Baser, K. H. C.; Haskologlu, I. C.; Erdag, E. Molecular Links Between Circadian Rhythm Disruption, Melatonin, and Neurodegenerative Diseases: An Updated Review. Molecules 2025, 30 (9). DOI: 10.3390/molecules30091888 From NLM.

(11) Meléndez-Fernández, O. H.; Liu, J. A.; Nelson, R. J. Circadian Rhythms Disrupted by Light at Night and Mistimed Food Intake Alter Hormonal Rhythms and Metabolism. Int J Mol Sci 2023, 24 (4). DOI: 10.3390/ijms24043392 From NLM.

(12) Sumová, A.; Sládek, M. Circadian Disruption as a Risk Factor for Development of Cardiovascular and Metabolic Disorders - From Animal Models to Human Population. Physiol Res 2024, 73 (Suppl 1), S321–s334. DOI: 10.33549/physiolres.935304 From NLM.

(13) Andreadi, A.; Andreadi, S.; Todaro, F.; Ippoliti, L.; Bellia, A.; Magrini, A.; Chrousos, G. P.; Lauro, D. Modified Cortisol Circadian Rhythm: The Hidden Toll of Night-Shift Work. Int J Mol Sci 2025, 26 (5). DOI: 10.3390/ijms26052090 From NLM.

(14) Ono, M.; Dai, Y.; Fujiwara, T.; Fujiwara, H.; Daikoku, T.; Ando, H.; Kuji, N.; Nishi, H. Influence of lifestyle and the circadian clock on reproduction. Reprod Med Biol 2025, 24 (1), e12641. DOI: 10.1002/rmb2.12641 From NLM.

(15) Yaw, A. M.; McLane-Svoboda, A. K.; Hoffmann, H. M. Shiftwork and Light at Night Negatively Impact Molecular and Endocrine Timekeeping in the Female Reproductive Axis in Humans and Rodents. Int J Mol Sci 2020, 22 (1). DOI: 10.3390/ijms22010324 From NLM.

(16) Ning, G.; Li, B. N.; Wu, H.; Shi, R. B.; Peng, A. J.; Wang, H. Y.; Zhou, X. Regulation of testosterone synthesis by circadian clock genes and its research progress in male diseases. Asian J Androl 2025, 27 (5), 564–573. DOI: 10.4103/aja20258 From NLM.

(17) Miyatake, A.; Morimoto, Y.; Oishi, T.; Hanasaki, N.; Sugita, Y.; Iijima, S.; Teshima, Y.; Hishikawa, Y.; Yamamura, Y. Circadian rhythm of serum testosterone and its relation to sleep: comparison with the variation in serum luteinizing hormone, prolactin, and cortisol in normal men. J Clin Endocrinol Metab 1980, 51 (6), 1365–1371. DOI: 10.1210/jcem-51-6-1365 From NLM.

(18) Li, L.; Lin, W.; Wang, Z.; Huang, R.; Xia, H.; Li, Z.; Deng, J.; Ye, T.; Huang, Y.; Yang, Y. Hormone Regulation in Testicular Development and Function. Int J Mol Sci 2024, 25 (11). DOI: 10.3390/ijms25115805 From NLM.

(19) Wang, Y.; Ni, C.; Li, X.; Lin, Z.; Zhu, Q.; Li, L.; Ge, R. S. Phthalate-Induced Fetal Leydig Cell Dysfunction Mediates Male Reproductive Tract Anomalies. Front Pharmacol 2019, 10, 1309. DOI: 10.3389/fphar.2019.01309 From NLM.

(20) Zhang, X. D.; Sun, J.; Zheng, X. M.; Zhang, J.; Tan, L. L.; Fan, L. L.; Luo, Y. X.; Hu, Y. F.; Xu, S. D.; Zhou, H.;, et al. Plin4 exacerbates cadmium-decreased testosterone level via inducing ferroptosis in testicular Leydig cells. Redox Biol 2024, 76, 103312. DOI: 10.1016/j.redox.2024.103312 From NLM.

(21) Lotti, F.; Maggi, M. Sexual dysfunction and male infertility. Nat Rev Urol 2018, 15 (5), 287–307. DOI: 10.1038/nrurol.2018.20 From NLM.

(22) Barbouni, K.; Jotautis, V.; Metallinou, D.; Diamanti, A.; Orovou, E.; Liepinaitienė, A.; Nikolaidis, P.; Karampas, G.; Sarantaki, A. When Weight Matters: How Obesity Impacts Reproductive Health and Pregnancy-A Systematic Review. Curr Obes Rep 2025, 14 (1), 37. DOI: 10.1007/s13679-025-00629-9 From NLM.

(23) Su, X.; Lin, D.; Luo, D.; Sun, M.; Wang, X.; Ye, J.; Zhang, M.; Zhang, Y.; Xu, X.; Yu, C.;, et al. Cyclophilin D participates in the inhibitory effect of high-fat diet on the expression of steroidogenic acute regulatory protein. J Cell Mol Med 2019, 23 (10), 6859–6871. DOI: 10.1111/jcmm.14569 From NLM.

(24) Wang, P.; Li, Q.; Wu, L.; Yu, X.; Zheng, Y.; Liu, J.; Yao, J.; Liu, Z.; Fan, S.; Li, Y. Association between the weight-adjusted-waist index and testosterone deficiency in adult males: a cross-sectional study. Sci Rep 2024, 14 (1), 25574. DOI: 10.1038/s41598-024-76574-9 From NLM. Liu, D.; Li, Y.; Ji, N.; Xia, W.; Zhang, B.; Feng, X. Association between weight-adjusted waist index and testosterone deficiency in adult American men: findings from the national health and nutrition examination survey 2013-2016. BMC Public Health 2024, 24 (1), 1683. DOI: 10.1186/s12889-024-19202-5 From NLM.

(25) Li, X.; Wen, Z.; Wang, Y.; Mo, J.; Zhong, Y.; Ge, R. S. Bisphenols and Leydig Cell Development and Function. Front Endocrinol (Lausanne) 2020, 11, 447. DOI: 10.3389/fendo.2020.00447 From NLM.

(26) Yang, X.; Lan, M.; Yang, J.; Xia, Y.; Han, L.; Zhang, L.; Fang, Y. Targeting Modifiable Risks: Molecular Mechanisms and Population Burden of Lifestyle Factors on Male Genitourinary Health. Int J Mol Sci 2025, 26 (19). DOI: 10.3390/ijms26199698 From NLM.

(27) Sudlow, C.; Gallacher, J.; Allen, N.; Beral, V.; Burton, P.; Danesh, J.; Downey, P.; Elliott, P.; Green, J.; Landray, M.;, et al. UK biobank: an open access resource for identifying the causes of a wide range of complex diseases of middle and old age. PLoS Med 2015, 12 (3), e1001779. DOI: 10.1371/journal.pmed.1001779 From NLM.

(28) Doherty, A.; Jackson, D.; Hammerla, N.; Plötz, T.; Olivier, P.; Granat, M. H.; White, T.; van Hees, V. T.; Trenell, M. I.; Owen, C. G.;, et al. Large Scale Population Assessment of Physical Activity Using Wrist Worn Accelerometers: The UK Biobank Study. PLoS One 2017, 12 (2), e0169649. DOI: 10.1371/journal.pone.0169649 From NLM.

(29) Fishbein, A. B.; Knutson, K. L.; Zee, P. C. Circadian disruption and human health. J Clin Invest 2021, 131 (19). DOI: 10.1172/jci148286 From NLM.

(30) Feng, H.; Yang, L.; Ai, S.; Liu, Y.; Zhang, W.; Lei, B.; Chen, J.; Liu, Y.; Chan, J. W. Y.; Chan, N. Y.;, et al. Association between accelerometer-measured amplitude of rest-activity rhythm and future health risk: a prospective cohort study of the UK Biobank. Lancet Healthy Longev 2023, 4 (5), e200–e210. DOI: 10.1016/s2666-7568(23)00056-9 From NLM.

(31) Cui, L.; Nie, X.; Guo, Y.; Ren, P.; Guo, Y.; Wang, X.; Li, R.; Hotaling, J. M.; Cairns, B. R.; Guo, J. Single-cell transcriptomic atlas of the human testis across the reproductive lifespan. Nat Aging 2025, 5 (4), 658–674. DOI: 10.1038/s43587-025-00824-2 From NLM.

(32) Bhasin, S.; Brito, J. P.; Cunningham, G. R.; Hayes, F. J.; Hodis, H. N.; Matsumoto, A. M.; Snyder, P. J.; Swerdloff, R. S.; Wu, F. C.; Yialamas, M. A. Testosterone Therapy in Men With Hypogonadism: An Endocrine Society Clinical Practice Guideline. J Clin Endocrinol Metab 2018, 103 (5), 1715–1744. DOI: 10.1210/jc.2018-00229 From NLM.

(33) Arlinghaus, A.; Bohle, P.; Iskra-Golec, I.; Jansen, N.; Jay, S.; Rotenberg, L. Working Time Society consensus statements: Evidence-based effects of shift work and non-standard working hours on workers, family and community. Ind Health 2019, 57 (2), 184–200. DOI: 10.2486/indhealth.SW-4 From NLM.

(34) Mulhall, J. P.; Trost, L. W.; Brannigan, R. E.; Kurtz, E. G.; Redmon, J. B.; Chiles, K. A.; Lightner, D. J.; Miner, M. M.; Murad, M. H.; Nelson, C. J.;, et al. Evaluation and Management of Testosterone Deficiency: AUA Guideline. J Urol 2018, 200 (2), 423–432. DOI: 10.1016/j.juro.2018.03.115 From NLM.

(35) Travicic, D. Z.; Becin, A. P.; Lalosevic, D.; Andric, S. A.; Kostic, T. S. Environmental light deprivation disrupts Leydig cell maturation and male reproductive development in rats. Sci Rep 2025, 15 (1), 37326. DOI: 10.1038/s41598-025-21236-7 From NLM.

(36) Liu, P. Y.; Takahashi, P. Y.; Yang, R. J.; Iranmanesh, A.; Veldhuis, J. D. Age and time-of-day differences in the hypothalamo-pituitary-testicular, and adrenal, response to total overnight sleep deprivation. Sleep 2020, 43 (7). DOI: 10.1093/sleep/zsaa008 From NLM.

(37) Liu, P. Y.; Reddy, R. T. Sleep, testosterone and cortisol balance, and ageing men. Rev Endocr Metab Disord 2022, 23 (6), 1323–1339. DOI: 10.1007/s11154-022-09755-4 From NLM.

(38) Bracci, M.; Zingaretti, L.; Martelli, M.; Lazzarini, R.; Salvio, G.; Amati, M.; Milinkovic, M.; Ulissi, A.; Medori, A. R.; Vitale, E.;, et al. Alterations in Pregnenolone and Testosterone Levels in Male Shift Workers. Int J Environ Res Public Health 2023, 20 (4). DOI: 10.3390/ijerph20043195 From NLM.

(39) de Souza, K. A.; Jackson, M.; Chen, J.; Reyes, J.; Muayad, J.; Tran, E.; Jackson, W.; Newell-Rogers, M. K.; Earnest, D. J. Shift work schedules alter immune cell regulation and accelerate cognitive impairment during aging. J Neuroinflammation 2025, 22 (1), 4. DOI: 10.1186/s12974-024-03324-z From NLM.

(40) Travicic, D. Z.; Miljkovic, D.; Andric, S. A.; Kostic, T. S. Circadian disruption impairs Leydig cell maturation and reproductive development in male rats. Reprod Biol Endocrinol 2025, 23 (1), 104. DOI: 10.1186/s12958-025-01440-w From NLM.

(41) Xu, Y.; Wang, L.; Cao, S.; Hu, R.; Liu, R.; Hua, K.; Guo, Z.; Di, H. J.; Hu, Z. Genipin improves reproductive health problems caused by circadian disruption in male mice. Reprod Biol Endocrinol 2020, 18 (1), 122. DOI: 10.1186/s12958-020-00679-9 From NLM.

(42) Liu, Q.; Wang, H.; Wang, H.; Li, N.; He, R.; Liu, Z. Per1/Per2 Disruption Reduces Testosterone Synthesis and Impairs Fertility in Elderly Male Mice. Int J Mol Sci 2022, 23 (13). DOI: 10.3390/ijms23137399 From NLM.

(43) Fusco, F.; Longo, N.; De Sio, M.; Arcaniolo, D.; Celentano, G.; Capece, M.; La Rocca, R.; Mangiapia, F.; Califano, G.; Morra, S.;, et al. Impact of Circadian Desynchrony on Spermatogenesis: A Mini Review. Front Endocrinol (Lausanne) 2021, 12, 800693. DOI: 10.3389/fendo.2021.800693 From NLM.

(44) Liu, T.; He, W.; Zhong, Z.; Lu, C.; Wu, L.; Wang, Z.; Smith, W. K.; Shi, Q.; Long, Q.; Wang, H. The circadian clock orchestrates spermatogonial differentiation and fertilization by regulating retinoic acid signaling in vertebrates. Natl Sci Rev 2025, 12 (3), nwae456. DOI: 10.1093/nsr/nwae456 From NLM.

(45) Li, S. Y.; Kumar, S.; Gu, X.; DeFalco, T. Testicular immunity. Mol Aspects Med 2024, 100, 101323. DOI: 10.1016/j.mam.2024.101323 From NLM.

(46) Kitchen, G. B.; Cunningham, P. S.; Poolman, T. M.; Iqbal, M.; Maidstone, R.; Baxter, M.; Bagnall, J.; Begley, N.; Saer, B.; Hussell, T.;, et al. The clock gene Bmal1 inhibits macrophage motility, phagocytosis, and impairs defense against pneumonia. Proc Natl Acad Sci U S A 2020, 117 (3), 1543–1551. DOI: 10.1073/pnas.1915932117 From NLM.

(47) Albrecht, U.; Ripperger, J. A. Circadian Clocks and Sleep: Impact of Rhythmic Metabolism and Waste Clearance on the Brain. Trends Neurosci 2018, 41 (10), 677–688. DOI: 10.1016/j.tins.2018.07.007 From NLM.

(48) Hedger, M. P. Immunophysiology and pathology of inflammation in the testis and epididymis. J Androl 2011, 32 (6), 625–640. DOI: 10.2164/jandrol.111.012989 From NLM.

(49) Jiang, S.; Li, H.; Zhang, L.; Mu, W.; Zhang, Y.; Chen, T.; Wu, J.; Tang, H.; Zheng, S.; Liu, Y.;, et al. Generic Diagramming Platform (GDP): a comprehensive database of high-quality biomedical graphics. Nucleic Acids Res 2025, 53 (D1), D1670–d1676. DOI: 10.1093/nar/gkae973 From NLM.

