## Supplementary materials for "Circadian Disruption Induces Testosterone Decline and Male Reproductive Impairment: Evidence from Epidemiological Studies and Animal Experiments"

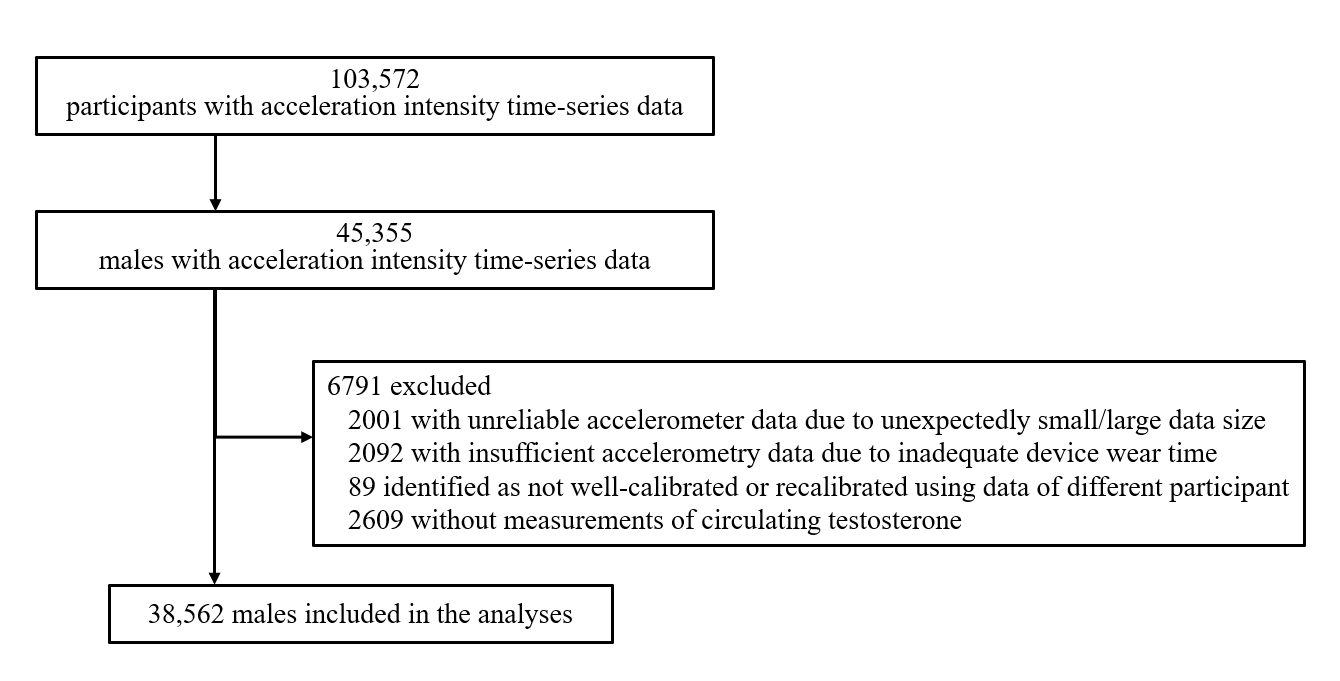

Supplementary Figure 1. Flow diagram of participant inclusion and exclusion.

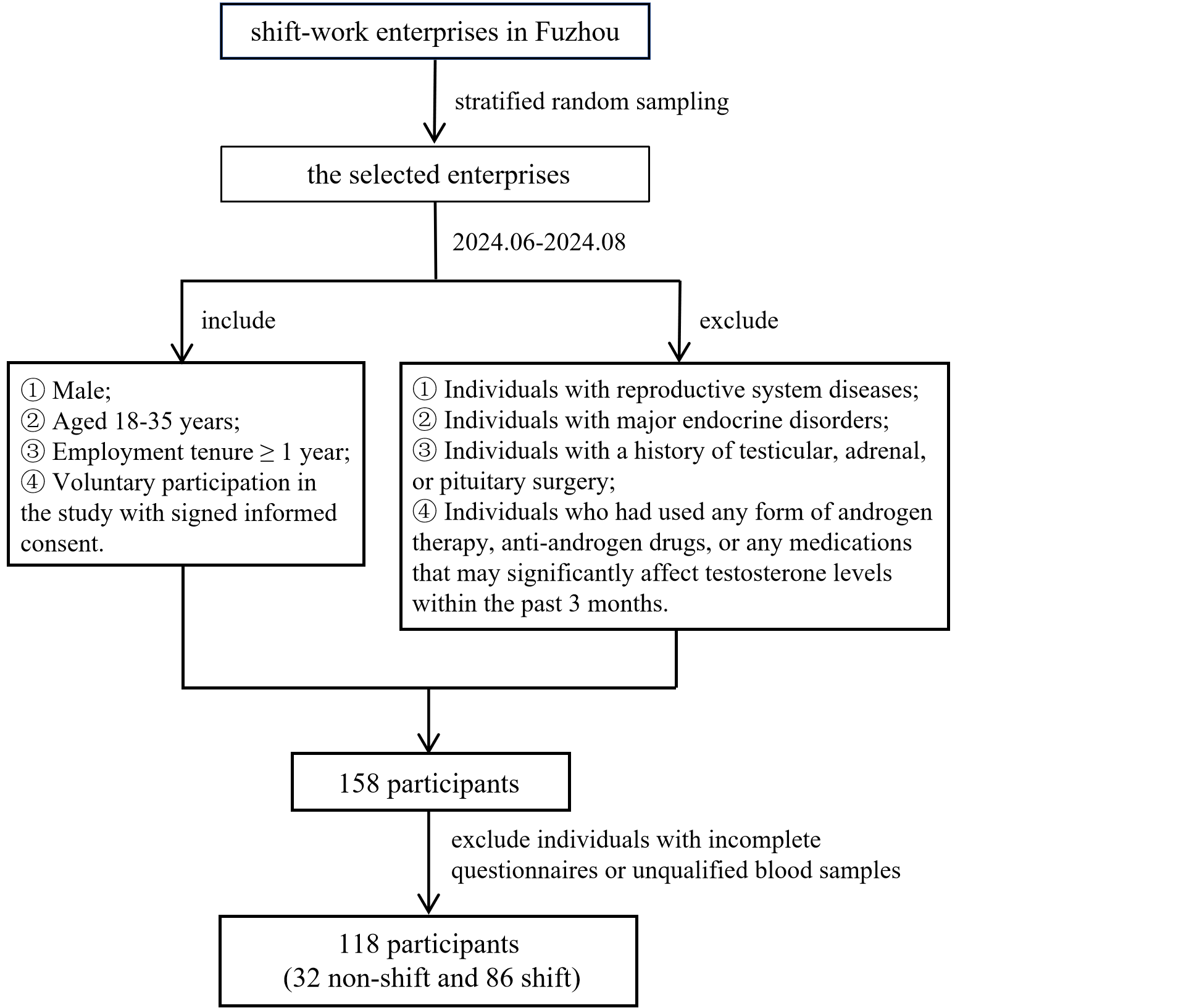

Supplementary Figure 2. Flow diagram of Fuzhou shift-work participant inclusion and exclusion.

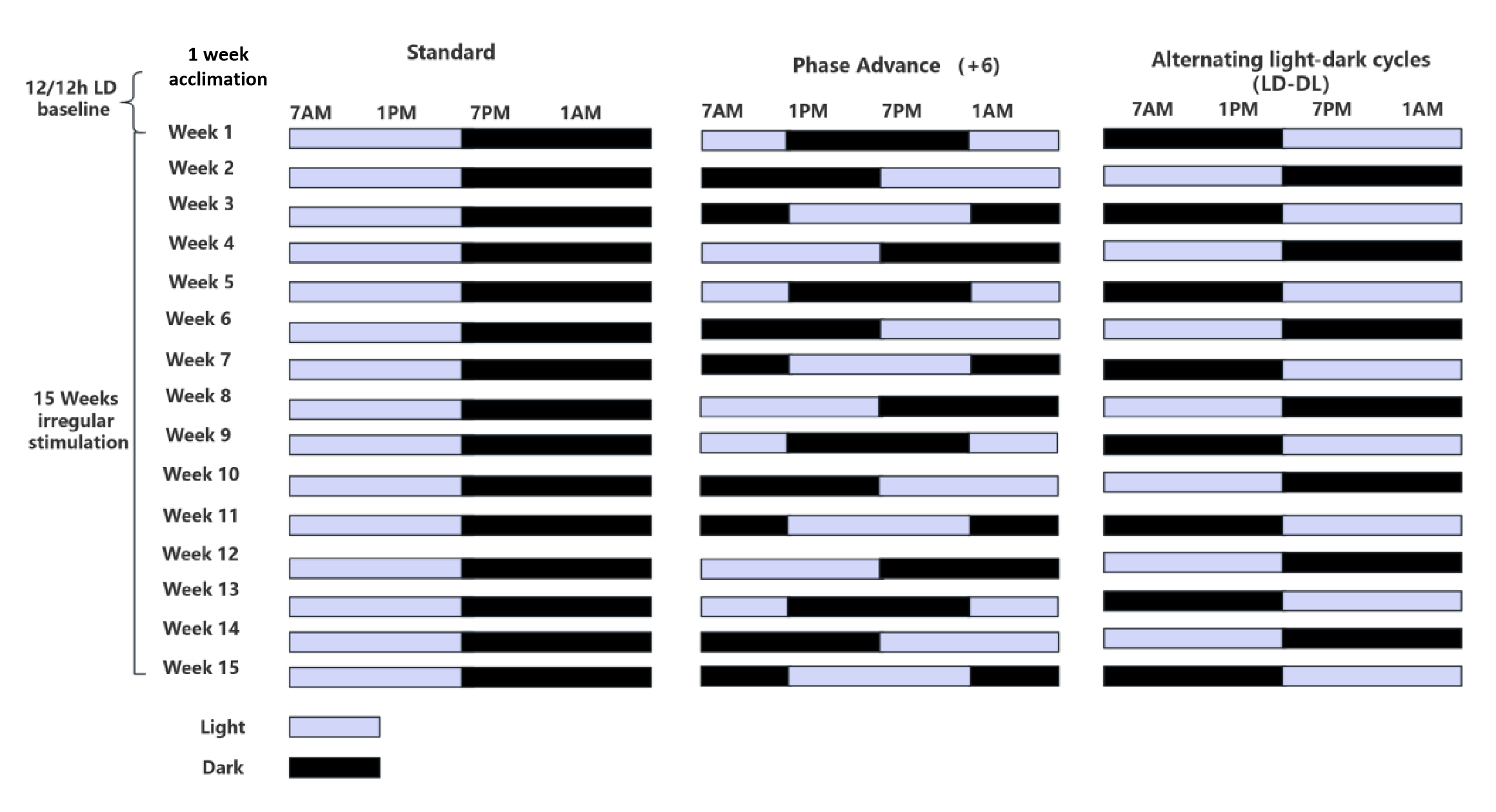

Supplementary Figure 3. C57BL/6 mouse circadian rhythm disruption model.
